# Temporal variations in international air travel: implications for modelling the spread of infectious diseases

**DOI:** 10.1101/2024.02.12.24302682

**Authors:** Jack Wardle, Sangeeta Bhatia, Anne Cori, Pierre Nouvellet

## Abstract

**Background:** The international flight network creates multiple routes by which pathogens can quickly spread across the globe. In the early stages of infectious disease outbreaks, analyses using flight passenger data to identify countries at risk of importing the pathogen are common and can help inform disease control efforts. A challenge faced in this modelling is that the latest aviation statistics (referred to as contemporary data) are typically not immediately available. Therefore, flight patterns from a previous year are often used (referred to as historical data). We explored the suitability of historical data for predicting the spatial spread of emerging epidemics.

**Methods:** We analysed monthly flight passenger data from the International Air Transport Association to assess how baseline air travel patterns were affected in outbreaks of MERS, Zika, and SARS-CoV-2 over the past decade. We then used a stochastic discrete time SEIR metapopulation model to simulate global spread of different pathogens, comparing how epidemic dynamics differed in simulations based on historical and contemporary data.

**Results:** We observed local, short-term disruptions to air travel from South Korea and Brazil for the MERS and Zika outbreaks we studied, whereas global and longer-term flight disruption occurred during the SARS-CoV-2 pandemic.

For outbreak events that were accompanied by local, small, and short-term changes in air travel, epidemic models using historical flight data gave similar projections of timing and locations of disease spread as when using contemporary flight data. However, historical data were less reliable to model the spread of an atypical outbreak such as SARS-CoV-2 in which there were durable and extensive levels of global travel disruption.

**Conclusions:** The use of historical flight data as a proxy in epidemic models is an acceptable practice except in rare, large epidemics that lead to substantial disruptions to international travel.

## INTRODUCTION

Localised outbreaks of emerging and re-emerging pathogens are often followed by international spread to multiple countries and continents (1, 2), with human population movement one of the key factors facilitating this spread. The international flight network plays a part in this, connecting populations separated by large distances with short travel times. Understanding the volume and spatiotemporal patterns of flight passengers can therefore provide insights into the routes by which a pathogen can spread (1, 2).

Analyses using flight passenger volumes have answered critical questions in the early phases of previous infectious disease epidemics. Early in the severe acute respiratory syndrome coronavirus-2 (SARS-CoV-2) pandemic, passenger data helped to identify the likely locations where the virus could be exported, assess the potential for travel restrictions to control spread, and estimate the true epidemic size in Wuhan based on cases identified among travellers to other countries (3–5). Similar studies were conducted for Ebola in West Africa (6, 7), and Zika (8, 9) and Yellow Fever (10) in the Americas.

Such studies can help control the spread of emerging epidemics through rapid communication of conclusions, increasing international awareness and aiding preparedness, surveillance, and response planning (5, 11, 12). As an outbreak unfolds in real-time, one challenge for spatiotemporal epidemic modelling is that current aviation statistics (referred to as ‘contemporary’ data throughout) are typically not immediately available. However, waiting for the data is not feasible in a rapidly growing epidemic. In addition, flight datasets are typically expensive to purchase. Consequently, the movement data used in models are often selected based on what is available, i.e. typically flight data from previous years.

For example, many of the studies evaluating the potential international spread of SARS-CoV-2 from China in early 2020 used passenger numbers from the corresponding months in 2019 (3, 4, 13–17), or occasionally 2018 (11, 18, 19). In a brief literature search of spatial epidemic models for SARS-CoV-2 including flight data, we found that only one of 10 studies attempted to characterise the actual 2020 flight patterns. That study scaled 2019 passenger data according to more up-to-date information on the numbers of planes departing from China (relative to the equivalent period in the year before) (14). The lack of up-to-date movement data is not unique to SARS-CoV-2 modelling analyses. Historical flight data were also used to model the international spread of Ebola, Zika, and Yellow Fever outbreaks because of data availability issues (6–10).

However, to our knowledge, no study has assessed whether historical datasets are a suitable proxy for contemporary flight patterns when modelling epidemic spatial spread. This is important given that epidemics can affect volumes and spatiotemporal patterns of travel due to public perception of risks or travel bans. In this paper, we explore the suitability of historical datasets for predicting the spatial spread of emerging epidemics. We assess whether implicit assumptions of consistent travel patterns over time are valid and their impact on key outputs of spatial models of infectious disease spread. We aim to: i) identify the extent to which flight volumes were disrupted by previous epidemics; ii) assess whether the most popular destinations for travellers from a given country changed over time and during epidemics; iii) simulate epidemics to compare epidemic model outputs when using historical versus contemporary movement data.

## METHODS

An overview of the methods is provided below; further details of the methodology are available in **Supplementary Section 1**.

We focus on three past epidemics to explore our aims: a Middle East respiratory syndrome (MERS) outbreak in South Korea from May to July 2015 involving 186 reported cases with 38 deaths (20); the Zika epidemic in Brazil that was declared a Public Health Emergency of International Concern (PHEIC) in February 2016 (21); and the SARS-CoV-2 pandemic which emerged in China at the end of 2019 (22).

### Flight passenger data

We used flight passenger data purchased from the International Air Transport Association (IATA) (23). The dataset contained the numbers of passengers that travelled between pairs of international airports each month from January 2012 to December 2020, which we aggregated to a country level.

To identify differences in passenger volumes during epidemics, we examined the monthly number of passengers departing from South Korea, Brazil, and China, and calculated changes in passenger numbers during the relevant epidemic period (defined in **Supplementary Section 1.i**) relative to the same month in the previous year. We also analysed the temporal variation in the flight destinations from each of these three countries, considered as the epidemic centres. We compared how the top 10 destinations (by monthly passenger volume) from the epidemic centres varied for a specified calendar month across the years 2012-2020 (we analysed the months at the beginning of the contemporary periods, see **Table S1**).

### Epidemics simulation study

We conducted a simulation study to compare the characteristics of epidemics modelled using “historical” flight passenger data from the year before the disease emerged with models that used “contemporary” flight data from the epidemic period.

### Epidemic model

We used a stochastic discrete time SEIR metapopulation model to simulate the global spread of a pathogen emerging in a single country, with the probability of movement between countries being informed by the IATA passenger data.

### Simulation scenarios

We simulated epidemics for three flight scenarios that used data corresponding to the MERS, Zika, and SARS-CoV-2 epidemic periods. The models used either contemporary or historical passenger data. The contemporary and historical periods for each flight scenario are defined in **Table S1**.

Across all flight scenarios, we simulated epidemics of pathogens with natural history parameter values similar to MERS, Zika, and SARS-CoV-2 (**Table S2**). These examples explored different basic reproduction numbers (R0, the average number of secondary cases generate by a primary case in a susceptible population) and generation times (time between infection of a case and their infector). Simulations were initiated with 100 infectious cases in the epidemic centre (South Korea, Brazil, and China for MERS, Zika, and SARS-CoV-2 flight scenarios respectively), ran for one year, and assumed that the global population was initially fully susceptible to infection. For each natural history, we simulated 100 epidemics with contemporary flight data and 100 epidemics with historical data. Combining the flight and natural history scenarios gave nine overall scenarios in which we compared historical and contemporary flight data.

For each simulated epidemic we computed the following metrics:

- **Number of invaded countries over time**: the number of countries with at least 10 cumulative infections at each day.
- **Invasion time in *i***: the time to country *i* experiencing its 10^th^ cumulative infection.

For the historical and contemporary simulations in each scenario, we summarised the distributions of each metric across all 100 simulations using the median, 2.5% and 97.5% quantiles. We ordered countries by their median invasion times to obtain the average **invasion ranking**. We identified the first *n* countries that were invaded with the contemporary flight data, and then calculated the percentage of those countries that were also invaded first when using historical data.

For the simulations using SARS-CoV-2 flight data and natural history, we used the invasion rankings to validate the performance of our model against independent case data from the World Health Organisation for the SARS-CoV-2 pandemic (24). We compared the first 10 countries to report 10 SARS-COV-2 cases (24) with the top 10 invasion rankings from our simulations. Simulations in this validation step were seeded in China in January 2020.

## RESULTS

The number of flight passengers departing South Korea and Brazil showed an increasing trend over time (especially pronounced in South Korea), with some within-year seasonal variation (**Figure 1A-B**). However, epidemic events in those countries were accompanied by deviations from long-term passenger trends. The numbers of people flying from South Korea in the months after the MERS epidemic started (June-August 2015) were between 6.5% and 16.2% lower than the equivalent months in 2014 (**Figure 1A**). Similarly, passenger departures from Brazil in the months after the declaration of Zika virus as a PHEIC (March-July 2016) were between 3.3% and 10.2% lower than the previous year (**Figure 1B**). June 2016 had the fourth lowest monthly passenger departures between January 2012 and February 2020, with the months with fewer departures all occurring in 2012. South Korea and Brazil, as well as China (**Figure 1C**), experienced very large reductions in air travel during the SARS-CoV-2 pandemic. The largest reduction in monthly departures was in April 2020 when passenger numbers decreased by 98.6%, 97.9%, and 98.6% for South Korea, Brazil, and China respectively.

**Figure 1.**
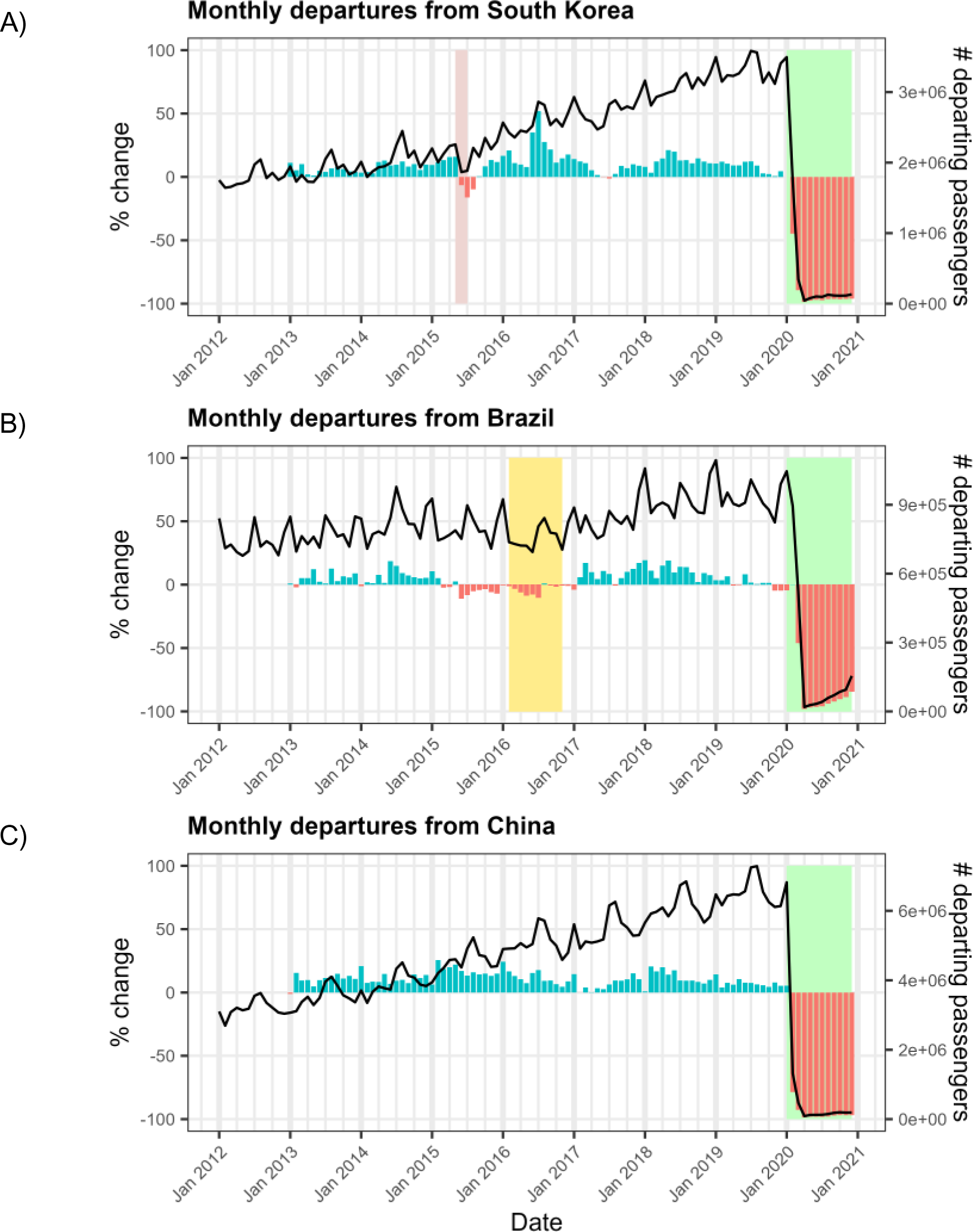
Changes in departing flight passenger volumes over time in A) South Korea, B) Brazil, and C) China. The black lines show the monthly numbers of flight passengers (right-hand axis) departing from South Korea, Brazil, and China from January 2012 to December 2020. The coloured bars denote the monthly percentage change in flight passenger numbers (left-hand axis) relative to the same calendar month in the previous year. Blue bars represent an increase in passenger numbers, red bars represent a decrease. Background coloured rectangles denote infectious disease outbreak periods (see **Supplementary Section 1.i**): Red = MERS; Yellow = Zika; Green = SARS-CoV-2.

The most popular destinations for flights from the three countries were generally consistent across years prior to the SARS-CoV-2 pandemic (**Figure 2**). Among these top 10s, there was some variation in the order, but in general the shifts in ordering were small. In South Korea and China, nine countries appeared consistently in the top 10 flight destinations for each year from 2012-2019. Brazil experienced more variability, with only six countries consistently in the top 10 destinations over 2012-2019. However, there was very little change in top destinations during the Zika PHEIC: of the 10 top destinations in March 2015, nine remained in the list for March 2016 (when Zika was a PHEIC). In all three epidemic centres, the year-to-year changes in destination lists were greatest between 2019 and 2020, but still modest: each country had 2/10 new countries in the 2020 lists.

**Figure 2.**
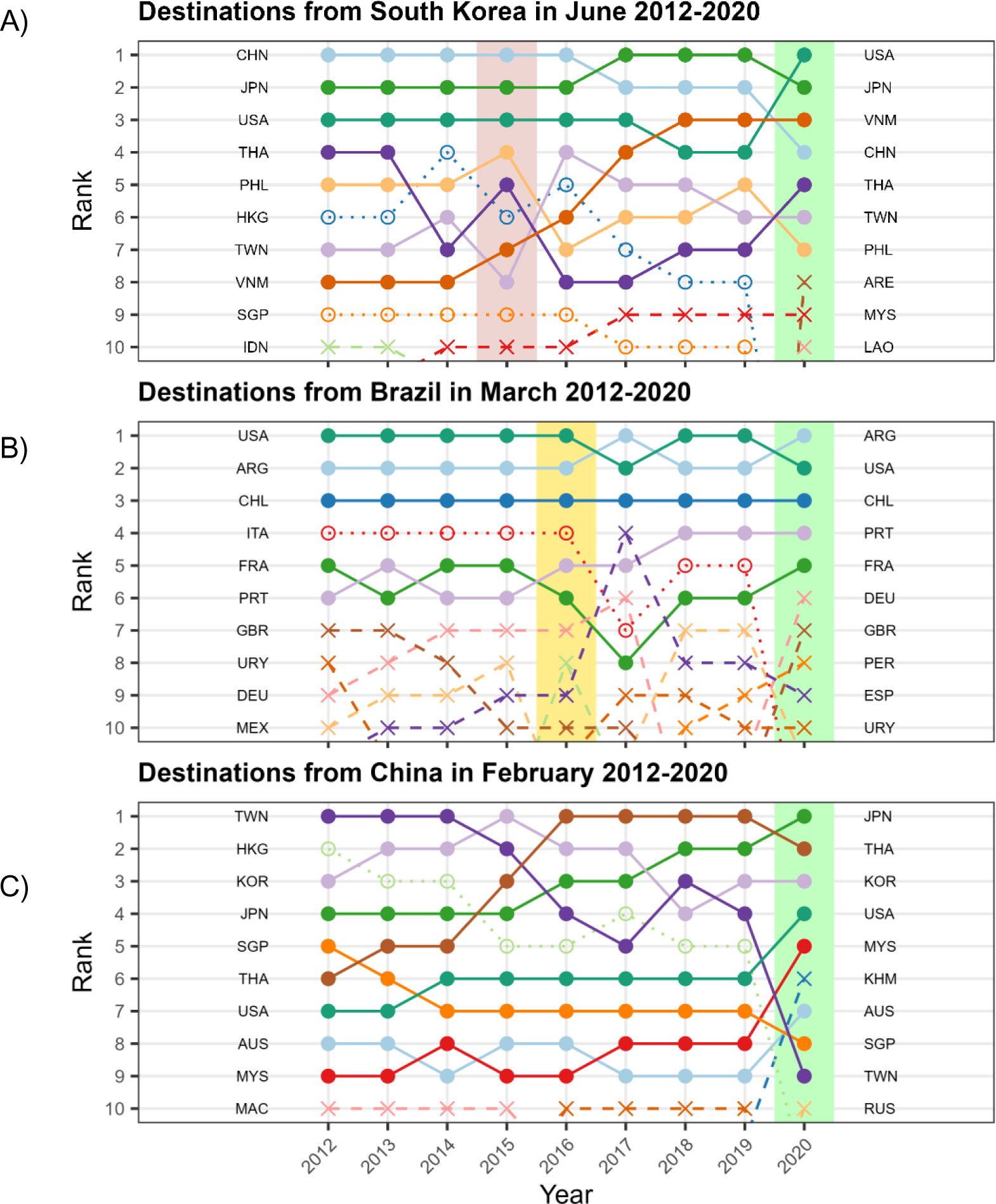
Top flight destinations over time in specific months for passengers departing from A) South Korea in June, B) Brazil in March, and C) China in February. The ranking is assigned using the total number of passengers departing by air in the specified month. The calendar month presented corresponds to the start of the periods shown in **Table S1** (i.e. early in the outbreak). Solid dots and lines are used for countries that feature in the top 10 destinations for a given country all years (2012 to 2020). Hollow circles and dotted lines represent countries that were in the top 10 destinations every year except 2020 (when there were extensive disruptions due to the SARS-CoV-2 pandemic). Crosses and dashed lines represent countries that were only in the top 10 for some years of the analysis period. Background coloured rectangles highlight the year in which epidemics occurred (red: MERS; yellow: Zika; green: SARS-CoV-2). <u>Country codes:</u> ARE: United Arab Emirates; ARG: Argentina; AUS: Australia; CHL: Chile; CHN: China; DEU: Germany; ESP: Spain; FRA: France; GBR: United Kingdom; HKG: Hong Kong; IDN: Indonesia; ITA: Italy; JPN: Japan; KHM: Cambodia; KOR: South Korea; LAO: Laos; MAC: Macau; MEX: Mexico; MYS: Malaysia; PER: Peru; PHL: Philippines; PRT: Portugal; RUS: Russia; SGP: Singapore; THA: Thailand; TWN: Taiwan; URY: Uruguay; USA: United States of America; VNM: Vietnam.

In simulated epidemics comparing historical and contemporary flight data from the MERS or Zika flight scenarios, we found very little difference in the rate the epidemics spread globally (**Figure 3**, second and third rows), irrespective of the pathogen natural histories. In contrast, in the SARS-CoV-2 flight scenario with extensive disruption to the global flight network, use of the historical flight data resulted in much earlier predicted spread than when using the contemporary flight data (**Figure 3**, first row). The differences were amplified by increasing the generation time or decreasing R0. In these SARS-CoV-2 flight scenario results, the difference in the median time to 50 countries being invaded was 25, 95, and 84 days for the SARS-CoV-2, MERS, and Zika natural history scenarios respectively. In all simulations, the SARS-CoV-2-like pathogen eventually spread to all countries, even with the extensive disruptions in the contemporary SARS-CoV-2 flight data. This was not the case when contemporary SARS-CoV-2 flight data was used for other natural history scenarios, which spread more slowly due to either longer generation times or smaller R0.

**Figure 3.**
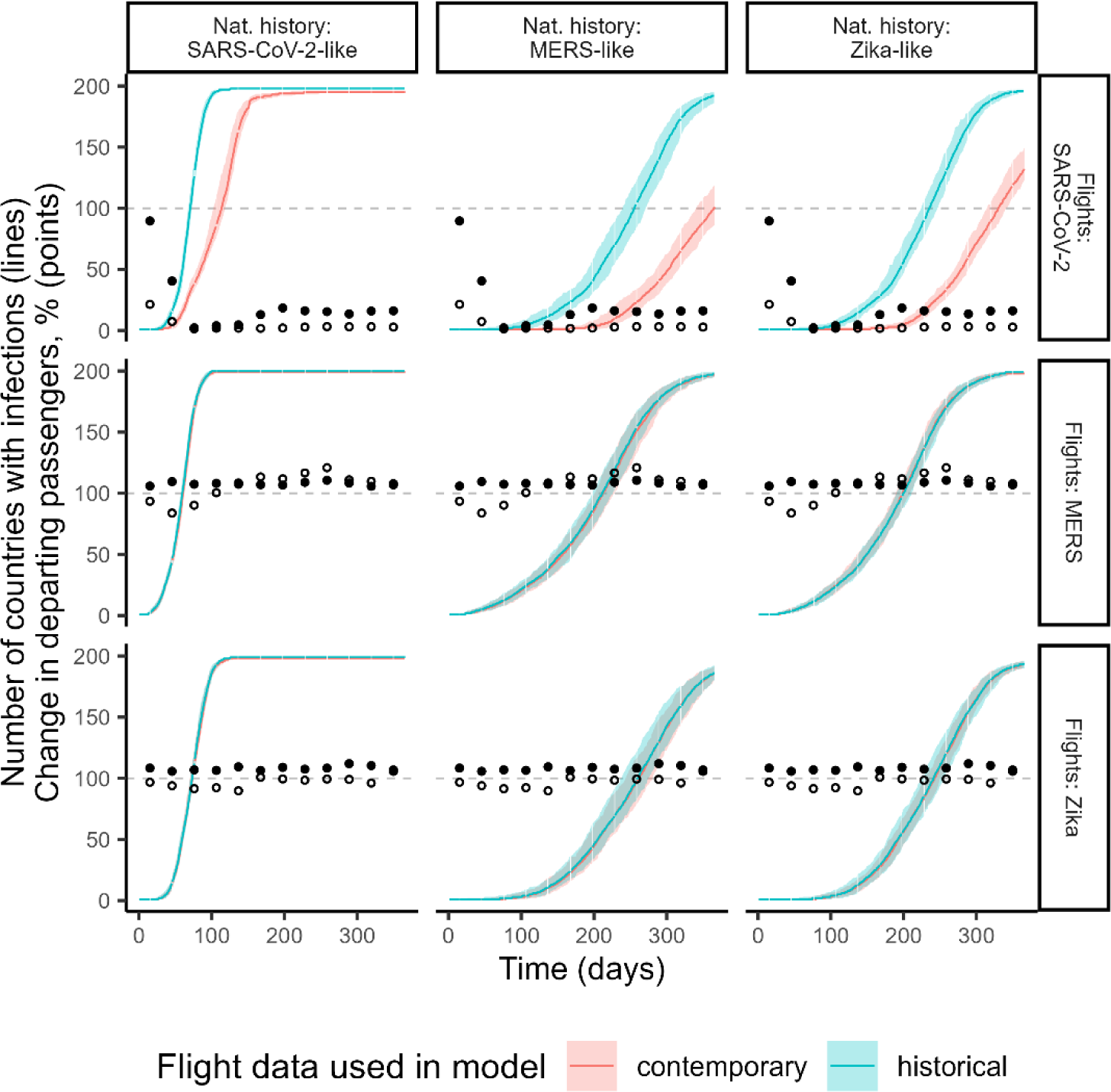
Number of countries invaded over time in epidemic simulations (lines) and relative percentage changes in departing passenger numbers (points). Rows correspond to different flight scenarios (**Table S1**) and columns to different natural history scenarios (**Table S2**). In each panel, the lines show the median number of countries with at least 10 cumulative infections across all 100 simulations for that flight data and natural history scenario combination. The coloured shading represents the 2.5% and 97.5% quantiles. Blue denotes simulation results using historical flight data, while red shows the simulations using contemporary flight data. Hollow and solid points display the percentage difference in the number of departing passengers from the epidemic centre or the difference from all countries respectively in each month of the contemporary period relative to the same month in the historical period (where a value of 100 corresponds to no change, 200 corresponds to a doubling, and 50 corresponds to a halving). The epidemic centre for the SARS-CoV-2, MERS and Zika flight scenarios are China, South Korea, and Brazil respectively. The travel volumes indicated by hollow circles influence how quickly the epidemic initially spreads out of the epidemic centre, while travel volumes shown by solid circles influence the rate of onward spread once the epidemic becomes established in a handful of other countries.

We explored how the differing invasion dynamics were influenced by the relative changes to the number of departing passengers in the contemporary versus historical data (**Figure 3**, circles). In the MERS and Zika flight scenarios, we found relatively small, short-term reductions to flight departures from the epidemic centre. This contrasted with a small increase in overall global flight departures, which reflected the trend of increasing flight volumes over time (**Figure S1**). The magnitude of the local changes seemed to have little impact on the initial spread from the epidemic centre and the subsequent rate of global epidemic spread. On the other hand, the SARS-CoV-2 flight scenario showed concurrent, large and durable reductions in both Chinese and total global flight departures.

Consequently, for the slower-growing MERS and Zika natural history scenarios, the epidemics remained localised at the epidemic centre until flight volumes from China recovered.

We found similar country invasion times when using contemporary and historical flight data for the MERS and Zika flight scenarios (**Figure 4**), across all three natural history scenarios. Differences in predicted invasion times were similar across countries invaded early and those invaded later in the epidemic (**Figure 4**). However, in the SARS-CoV-2 flight scenario, we found that using historical passenger data substantially underestimated the invasion times. Again, the invasion delay was amplified with larger generation times, with the median underestimation in invasion time ranging from 28 days (2.5% and 97.5% quantiles: 9, 55 days) to 93 days (54, 135 days) for the SARS-CoV-2 and Zika natural history scenarios. For the SARS-CoV-2 natural history scenario, the delays were more marked for countries invaded later, likely reflecting that early invasions occurred when there was less disruption to global travel, while the later countries to be invaded were in a period when there was increased disruption to passenger volumes (**Figure 2**, first row). Conversely, for the slower growing MERS and Zika natural history scenarios, differences in invasion were smaller for countries invaded later because their invasion occurred at times when there was relatively less disruption (compared to the period when the early countries were invaded) (**Figure 2**, first row).

**Figure 4.**
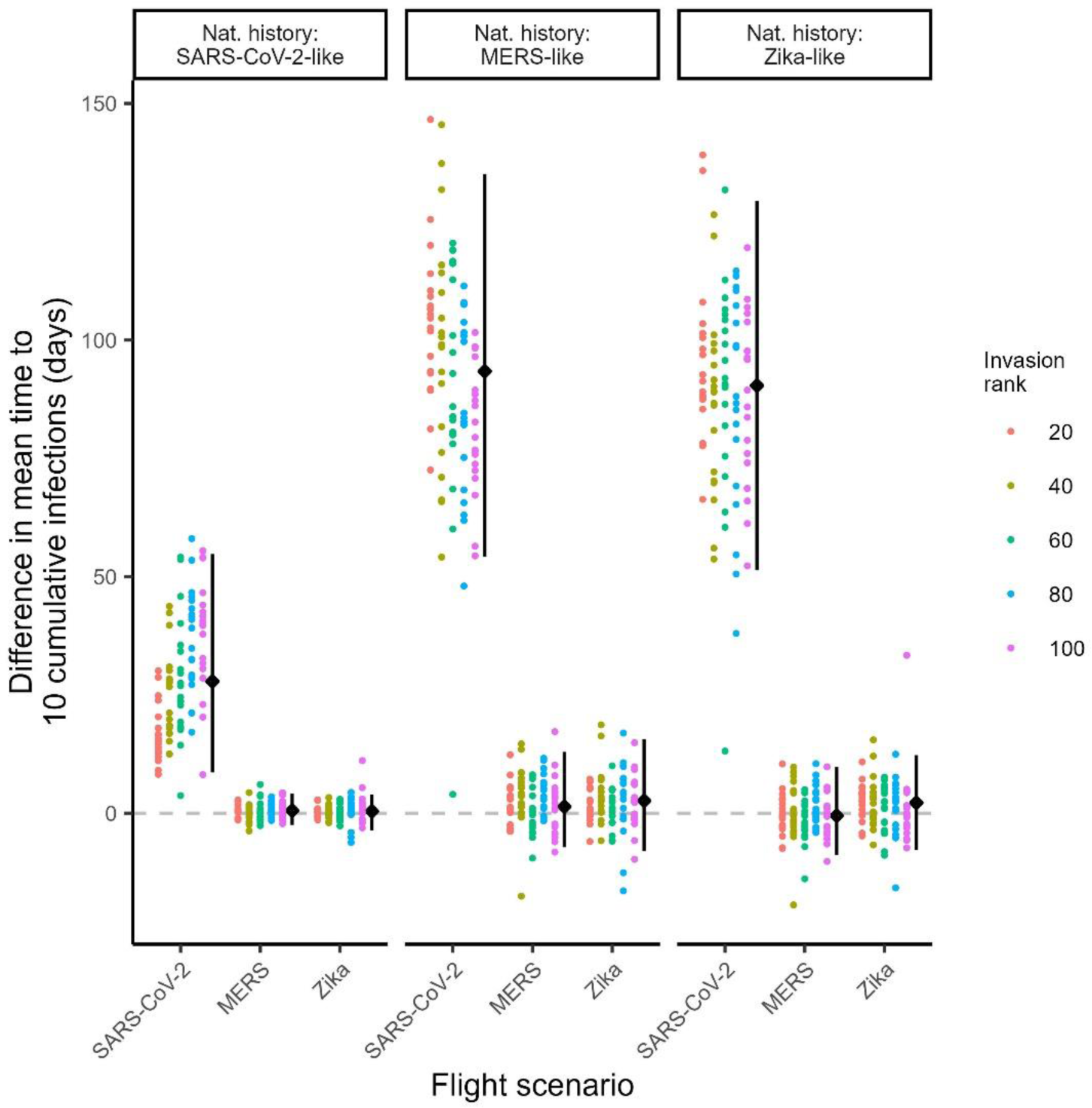
Difference in mean invasion times for simulations using contemporary versus historical flight data. The three panels correspond to different pathogen natural history scenarios. Within each panel, the x axis represents the three flight scenarios. Within each scenario combination, each dot represents a single country. The y-axis shows the mean invasion time using contemporary data minus the mean invasion time using historical data. Values above the dashed line means invasion was slower with the contemporary data. The colour of the dots shows the grouping of countries by the average rank in which they were invaded in the simulations using contemporary flight data. For example, the red invasion rank labelled ‘20’ corresponds to the first 20 countries to be invaded on average across simulations for that combination of natural history and flight scenarios. The black dots summarise the median difference in invasion time across the 100 countries shown, with the error bar showing the 2.5% and 97.5% quantiles.

Despite some underestimation of invasion times, there was generally good agreement in the first *n* invaded countries predicted using historical and contemporary flight data across all natural history scenarios (**Figure 5**). Across simulation scenarios, 60-100% of the first 10 countries invaded using historical flight data also featured in the first 10 countries invaded using contemporary data. This increased to 80-100% for the first 20 invaded countries.

**Figure 5.**
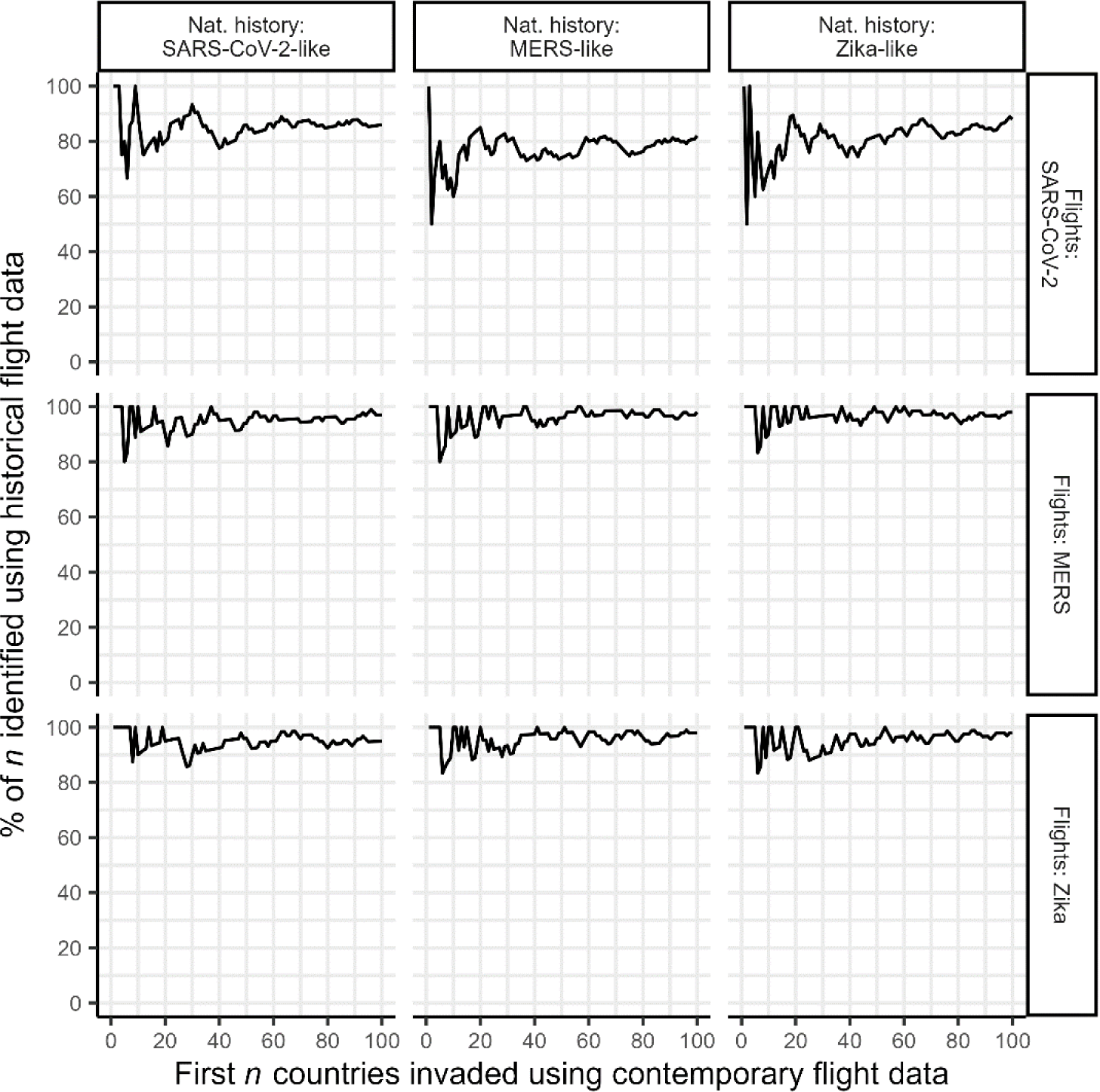
Similarity in invasion order for simulations using contemporary versus historical flight data. Rows correspond to different flight scenarios and columns to different natural histories. For each scenario, countries were ordered based on their median invasion times (defined as the time to a country experiencing the 10^th^ cumulative infection), to generate an average invasion ranking for both historical and contemporary simulations. The x-axis indicates the first *n* countries that were projected to be invaded in the contemporary flight data simulations. The y-axis shows the percentage of those countries that were also predicted by simulations based on historical data.

Since our findings are based on simulations, we assessed the extent to which predicted invasion orders reflected reality by comparing our model predictions to independent data on the early spatial spread of COVID-19. Our model performed well in predicting the early countries to report SARS-CoV-2 cases (**Table S3**). Seven of the first eight countries invaded in the model matched the first eight countries to report cases.

## DISCUSSION

Mathematical models of infectious disease spread relying on global flight data are often used in real-time to inform epidemic control efforts. Delayed publication of the latest flight passenger statistics means that models are often constrained to using historical data, typically from the previous year, and therefore do not capture changes to travel patterns and volumes that are caused by the outbreak. In this work, we showed that the standard practice of using historical data generally leads to similar projections of the timing and order of epidemic spread to other countries, compared to using contemporary flight data, for epidemic events with localized, relatively small, short-term mobility changes (such as those experienced during the MERS and Zika outbreaks). The consistency in the predicted order in which the epidemic reached countries is not surprising given our findings that the most common flight destinations were relatively stable over time.

Historical flight data were less suitable for modelling an atypical epidemic such as SARS-CoV-2 with durable, extensive levels of global travel disruption. Although the locations projected to be invaded early were consistent between historical and contemporary flight data, the projected invasion times were vastly underestimated when using historical data. This could lead to the dismissal of preventative interventions that are perceived as too slow for the projected speed of invasion (e.g. building emergency healthcare facilities). It may also reduce public trust in model outputs which could have implications such as decreasing compliance with interventions.

Our work suggests that correcting historical data to predict the contemporary spread of a pathogen would only be necessary for rare events with extensive travel disruptions. In such situations, a correction factor could be applied to historical flight data, as in the approach by Menkir et al (14). However, accurately predicting complex changes to travel in real-time is likely to be challenging.

While our study focused on the robustness of using historical flight data in real-time epidemic models, our findings also provide insights on the potential impact of travel restrictions. Our simulations suggest that large, widespread mobility reductions are needed to substantially impact disease spread. In the MERS and Zika flight scenarios, local, small, and short-term changes in mobility had little impact on the global spread of a pathogen. In the SARS-CoV-2 flight scenario, a rapid decrease in the number of departing passengers from the epidemic centre was soon followed by similar decreases globally. Although this substantially delayed the international spread of the epidemic in our simulations, ultimately all countries were still infected as international travel recovered, and eventually experienced similar epidemic sizes and peak sizes (**Figure S2**).

Therefore, travel restrictions seem to be insufficient to interrupt transmission sustainably but could provide an opportunity to prepare for the arrival of a pathogen. However, the substantial economic and political costs of introducing travel restrictions (25–28) mean that restrictions will only be worthwhile if the delay they generate is used sensibly, such as for the development of diagnostic, pharmaceutical, and non-pharmaceutical tools, and the logistics of their delivery.

The reductions in air travel in the contemporary SARS-CoV-2 flight data resulted in median delays to invasion of 25 days across the first 50 countries for the SARS-CoV-2 natural history scenario, rising to 95 days for the MERS natural history simulations. For context, over 1.3 million people were vaccinated by day 26 of the UK COVID-19 vaccination rollout, increasing to over 23.3 million by day 95 (29). Although these statistics do not account for the time to develop, manufacture, and distribute vaccines, they provide an example of the speed at which response measures can be implemented.

Our work is limited as our model has not been extensively validated against epidemiological data. However, validation of our SARS-CoV-2 scenarios found that the first countries invaded in our model generally matched the first countries to report cases in early 2020 (24). Further validation is challenging due to variability in the reporting of early cases across countries, with reporting potentially reflecting a country’s capacity to detect and report cases effectively, rather than their true burden.

Future research could investigate whether predictions of epidemic dynamics are improved by combining flight data with real-time movement indicators, e.g. changes in movement around airports from platforms such as Google or Meta (30, 31). However, overall, we showed that using historical instead of contemporary flight data had limited impact on simulated epidemic dynamics for two flight scenarios (MERS and Zika) and a range of pathogen natural histories. Only for the extreme SARS-CoV-2 flight scenario, with an almost complete shutdown of international travel, were projections of invasion times significantly underestimated. We note that the ability to use historical flight passenger data depends on scientists having access to these data; it is essential that those involved in epidemic response have timely access to data and access is not prevented by financial barriers.

## Data Availability

All code used in this analysis is available at https://github.com/j-wardle/flight_passenger_paper. Flight passenger data used in the analysis were purchased from IATA(https://www.iata.org).

## Funding acknowledgements

This study is partially funded by the National Institute for Health and Care Research (NIHR) Health Protection Research Unit in Modelling and Health Economics, a partnership between the UK Health Security Agency, Imperial College London and LSHTM (grant code NIHR200908); and acknowledges funding from the MRC Centre for Global Infectious Disease Analysis (reference MR/X020258/1), funded by the UK Medical Research Council (MRC). This UK funded award is carried out in the frame of the Global Health EDCTP3 Joint Undertaking; and acknowledges funding by Community Jameel. JW acknowledges research funding from the Wellcome Trust (grant 102169/Z/13/Z). AC was supported by the Academy of Medical Sciences Springboard scheme, funded by the AMS, Wellcome Trust, BEIS, the British Heart Foundation and Diabetes UK [REF:SBF005\1044]. Disclaimer: The views expressed are those of the author(s) and not necessarily those of the NIHR, UK Health Security Agency or the Department of Health and Social Care. The funders had no role in study design, data collection and analysis, decision to publish, or preparation of the manuscript.

## Author Contributions

JW: Conceptualization, Formal analysis, Methodology, Visualization, Writing – original draft, Writing – review and editing.

SB: Conceptualization, Formal analysis, Methodology, Writing – review and editing.

AC: Conceptualization, Formal analysis, Methodology, Supervision, Writing – review and editing.

PN: Conceptualization, Formal analysis, Methodology, Supervision, Writing – review and editing.

## REFERENCES

1. Tatem AJ, Rogers DJ, Hay SI. Global transport networks and infectious disease spread. Advances in parasitology. 2006;62:293–343.

2. Findlater A, Bogoch, II. Human Mobility and the Global Spread of Infectious Diseases: A Focus on Air Travel. Trends Parasitol. 2018;34(9):772–83.

3. Wu JT, Leung K, Leung GM. Nowcasting and forecasting the potential domestic and international spread of the 2019-nCoV outbreak originating in Wuhan, China: a modelling study. Lancet. 2020;395(10225):689–97.

4. Chinazzi M, Davis JT, Ajelli M, Gioannini C, Litvinova M, Merler S, et al. The effect of travel restrictions on the spread of the 2019 novel coronavirus (2019-nCoV) outbreak. medRxiv. 2020.

5. Imai N, Dorigatti I, Cori A, Riley S, Ferguson N. Estimating the potential total number of novel Coronavirus (2019-nCoV) cases in Wuhan City, China 2020 [17 January 2020]. 2020.

6. Read JM, Diggle PJ, Chirombo J, Solomon T, Baylis M. Effectiveness of screening for Ebola at airports. The Lancet. 2015;385(9962):23-4.

7. Bogoch II, Creatore MI, Cetron MS, Brownstein JS, Pesik N, Miniota J, et al. Assessment of the potential for international dissemination of Ebola virus via commercial air travel during the 2014 west African outbreak. The Lancet. 2015;385(9962):29-35.

8. Gardner LM, Bóta A, Gangavarapu K, Kraemer MUG, Grubaugh ND. Inferring the risk factors behind the geographical spread and transmission of Zika in the Americas. PLOS Neglected Tropical Diseases. 2018;12(1):e0006194.

9. Massad E, Tan S-H, Khan K, Wilder-Smith A. Estimated Zika virus importations to Europe by travellers from Brazil. Global Health Action. 2016;9(1):31669.

10. Dorigatti I, Morrison S, Donnelly CA, Garske T, Bowden S, Grills A. Risk of yellow fever virus importation into the United States from Brazil, outbreak years 2016–2017 and 2017–2018. Scientific Reports. 2019;9(1):20420.

11. De Salazar PM, Niehus R, Taylor A, Buckee C, Lipsitch M. Using predicted imports of 2019-nCoV cases to determine locations that may not be identifying all imported cases. medRxiv. 2020:2020.02.04.20020495.

12. Bhatia S, Imai N, Cuomo-Dannenburg G, Baguelin M, Boonyasiri A, Cori A, et al. Estimating the number of undetected COVID-19 cases among travellers from mainland China [version 3; peer review: 3 approved]. Wellcome Open Research. 2021;5(143).

13. Pullano G, Pinotti F, Valdano E, Boëlle PY, Poletto C, Colizza V. Novel coronavirus (2019-nCoV) early-stage importation risk to Europe, January 2020. Euro Surveill. 2020;25(4).

14. Menkir TF, Chin T, Hay J, Surface ED, De Salazar PM, Buckee CO, et al. Estimating the number of undetected COVID-19 cases exported internationally from all of China. medRxiv. 2020:2020.03.23.20038331.

15. Adiga A, Venkatramanan S, Schlitt J, Peddireddy A, Dickerman A, Bura A, et al. Evaluating the impact of international airline suspensions on the early global spread of COVID-19. medRxiv. 2020:2020.02.20.20025882.

16. Bogoch II, Watts A, Thomas-Bachli A, Huber C, Kraemer MUG, Khan K. Potential for global spread of a novel coronavirus from China. Journal of Travel Medicine. 2020;27(2).

17. Gilbert M, Pullano G, Pinotti F, Valdano E, Poletto C, Boëlle P-Y, et al. Preparedness and vulnerability of African countries against importations of COVID-19: a modelling study. The Lancet. 2020;395(10227):871–7.

18. Lai S, Bogoch II, Ruktanonchai NW, Watts A, Lu X, Yang W, et al. Assessing spread risk of Wuhan novel coronavirus within and beyond China, January-April 2020: a travel network-based modelling study. medRxiv. 2020:2020.02.04.20020479.

19. Bogoch II, Watts A, Thomas-Bachli A, Huber C, Kraemer MUG, Khan K. Pneumonia of unknown aetiology in Wuhan, China: potential for international spread via commercial air travel. Journal of Travel Medicine. 2020;27(2).

20. Arabi YM, Balkhy HH, Hayden FG, Bouchama A, Luke T, Baillie JK, et al. Middle East Respiratory Syndrome. N Engl J Med. 2017;376(6):584–94.

21. McCloskey B, Endericks T. The rise of Zika infection and microcephaly: what can we learn from a public health emergency? Public Health. 2017;150:87–92.

22. Hu B, Guo H, Zhou P, Shi Z-L. Characteristics of SARS-CoV-2 and COVID-19. Nature Reviews Microbiology. 2021;19(3):141–54.

23. International Air Transport Association (IATA). Flight passenger data available from: https://iata.org.

24. World Health Organisation. WHO Coronavirus (COVID-19) dashboard. Available at: https://data.who.int/dashboards/covid19/about. 2023.

25. Ferrell C, Agarwal P. Flight Bans and the Ebola Crisis Policy Recommendations for Future Global Health Epidemics. Harvard Public Health Review. 2018;14:1–14.

26. Iacus SM, Natale F, Santamaria C, Spyratos S, Vespe M. Estimating and projecting air passenger traffic during the COVID-19 coronavirus outbreak and its socio-economic impact. Safety Science. 2020;129:104791.

27. Nhamo G, Dube K, Chikodzi D. COVID-19 and Implications for the Aviation Sector: A Global Perspective. In: Nhamo G, Dube K, Chikodzi D, editors. Counting the Cost of COVID-19 on the Global Tourism Industry. Cham: Springer International Publishing; 2020. p. 89–107.

28. Seyfi S, Hall CM, Shabani B. COVID-19 and international travel restrictions: the geopolitics of health and tourism. Tourism Geographies. 2023;25(1):357–73.

29. UK Health Security Agency. Coronavirus (COVID-19) dashboard. Available at: https://coronavirus.data.gov.uk/details/download. 2021.

30. Google. COVID-19 Community Mobility Reports. Available at: https://www.google.com/covid19/mobility/. 2023.

31. Meta. Data for Good. Available at: https://dataforgood.facebook.com/dfg/covid-19. 2023.

