## Supplementary material for "Temporal variations in international air travel: implications for modelling the spread of infectious diseases"

### 1. Methods

#### i. *Flight passenger data*

We analysed flight passenger data purchased from the International Air Transport Association (IATA) (1). The dataset contained the numbers of passengers that travelled between pairs of international airports each month in the period January 2012 to December 2020. We aggregated passenger numbers to a country level to obtain the monthly numbers of people travelling by air from country *i* to country *j*. In processing the data, we considered only the origin and ultimate destination of the journey, ignoring any countries that were passed through due to connecting flights.

To identify differences in flight passenger volumes in epidemic periods relative to non-epidemic periods, we examined the monthly number of flight passengers departing from South Korea, Brazil, and China, and calculated the changes in passenger numbers during the relevant epidemic period relative to the same month in the previous year. The epidemic periods for each outbreak in this study were:

- **MERS**: May to July 2015;
- **Zika**: February to November 2016 (the period in which Zika was classed as a Public Health Emergency of International Concern);
- **SARS-CoV-2**: End of December 2019 to December 2020 (we analysed the first year of the pandemic).

Motivated by the common use of historical flight passenger data to model where a pathogen might spread, we also analysed the temporal variation in the flight destinations from each of the three countries, considered as the epidemic centres. We compared how the top 10 destinations (by monthly passenger volume) from the epidemic centres varied for a specified calendar month across the years 2012-2020. The months we selected were those at the beginning of the contemporary periods (see **Table S1)**.

#### ii. *Simulation study*

We conducted a simulation study to compare the characteristics of epidemics modelled using "historical" flight passenger data from the year before the disease emerged with models that used "contemporary" flight data from the epidemic period. Details of the historical and contemporary periods are shown in **Table S1**.

*a. Epidemic model*

We used a stochastic discrete time SEIR metapopulation model to simulate the global spread of a pathogen that emerges or re-emerges in one country. Each country formed one of *N* patches (*N*=200) in the model. In each patch there were susceptible (S), exposed (E), infected (I) and recovered (R) compartments. The probability of movement between different patches during day *t* was defined by a movement matrix, **P**, with entries $p^t_{i\to j}$. We generated **P** from the IATA data by computing the probability that a person in country *i* will travel to *j* on day *t*, calculated as:

$$p^t_{i\to j} = p^t_{i,fly}\frac{\phi^t_{i\to j}}{\sum_{x,x\neq i}\phi^t_{i\to x}}$$

where $p^t_{i,fly}$ is the probability that an individual in *i* takes an international flight on day *t*, $\phi^t_{i\to j}$ is the number of people flying from *i* to *j* on day *t*, and $\sum_{x,x\neq i}\phi^t_{i\to x}$ is the total number of people flying from *i* to all destinations on day *t*. We calculated $p^t_{i,fly}$ from the monthly aggregated IATA data by assuming passenger numbers were uniformly distributed across the month and dividing the number of international passenger flight departures on day *t* by $N_i$, the population size of country *i* (2). Our model therefore assumed that all people in a country were equally likely to fly.

For each day we modelled two processes in turn: 1) transitions between disease compartments; and 2) spatial movement. It was therefore possible that somebody progressed through disease stages and moved location in the same time step. The equations for the two processes in our spatial epidemic model at each time step (with *Δt* = 1 day) were therefore:

***Process 1: Disease progression***

$$S_i \overset{\Delta t}{\to} E_i \sim Binomial(S_i, 1 - \exp\,(-\frac{\beta I_i}{N_i} * \Delta t))$$

$$E_i \overset{\Delta t}{\to} I_i \sim Binomial(E_i, 1 - \exp(-\delta * \Delta t))$$

$$I_i \overset{\Delta t}{\to} R_i \sim Binomial(I_i, 1 - \exp\,(-\gamma * \Delta t))$$

***Process 2: Spatial movement***

$$(S_i \overset{\Delta t}{\to} S_1,\, S_i \overset{\Delta t}{\to} S_2, \dots,\, S_i \overset{\Delta t}{\to} S_N) \sim Multinomial(S_i,\, p^t_{i\to 1},\, p^t_{i\to 2}, \dots,\, p^t_{i\to N})$$

$$(E_i \xrightarrow{\Delta t} E_1,\ E_i \xrightarrow{\Delta t} E_2, \dots,\ E_i \xrightarrow{\Delta t} E_N) \sim Multinomial(E_i,\ p^t_{i\to1},\ p^t_{i\to2}, \dots,\ p^t_{i\to N})$$

$$(I_i \xrightarrow{\Delta t} I_1,\ I_i \xrightarrow{\Delta t} I_2, \dots,\ I_i \xrightarrow{\Delta t} I_N) \sim Multinomial(I_i,\ p^t_{i\to1},\ p^t_{i\to2}, \dots,\ p^t_{i\to N})$$

$$(R_i \xrightarrow{\Delta t} R_1,\ R_i \xrightarrow{\Delta t} R_2, \dots,\ R_i \xrightarrow{\Delta t} R_N) \sim Multinomial(R_i,\ p^t_{i\to1},\ p^t_{i\to2}, \dots,\ p^t_{i\to N})$$

where β is the per capita transmission rate, 1/δ and 1/γ are the mean latent and infectious periods respectively.

*b. Simulation scenarios*

We simulated epidemics for three flight scenarios that used data corresponding to the MERS, Zika, and SARS-CoV-2 epidemic periods. For each of these, we compared the dynamics of simulated epidemics when **P** was informed by contemporary flight data with simulations where **P** was informed by historical flight data; the periods we defined as contemporary and historical are detailed in **Table S1**.

For each of the flight data comparisons above, we simulated epidemics of pathogens with natural history parameter values similar to MERS, Zika, and SARS-CoV-2 (see **Table S2**) These examples explored different basic reproduction numbers (R0, the average number of secondary cases generate by a primary case in a susceptible population) and generation times (time between infection of a case and their infector). Although they were initially informed by the natural history parameter values of MERS, Zika and SARS-CoV-2 from the literature, some adjustments have been made to the parameters we used so that we could explore a range of transmissibility scenarios across our three examples. In addition, our model does not attempt to model the vectors required for transmission of Zika. Instead, the Zika-like simulations are used to illustrate the potential spread of a pathogen with a long generation time.

Combining the three flight scenarios and three natural history scenarios gave nine scenarios in which we compared historical and contemporary flight data. For each, we ran 100 simulated epidemics for one year, starting with 100 infectious cases of the pathogen in the epidemic centre (South Korea, Brazil, and China for MERS, Zika, and SARS-CoV-2 flight scenarios respectively) and assuming that the rest of the global population were fully susceptible (using national population data from the World Bank (2)).

We characterised each simulated epidemic with the following metrics:

- **Number of invaded countries over time**: the number of countries with at least 10 cumulative infections at each day.
- **Invasion time in *i***: the time to a country *i* experiencing its 10$^{th}$ cumulative infection.

For the historical and contemporary simulations in each scenario, we summarised the distributions of each metric across the 100 simulated epidemics using the median, 2.5% and 97.5% quantiles. We ordered countries by their median invasion times to obtain the average **invasion ranking**. We identified the first *n* countries that were invaded with the contemporary flight data, and then calculated the percentage of those countries that were also invaded first when using historical data.

For the simulations using SARS-CoV-2 flight data and natural history, we used the invasion rankings to validate the performance of the model against independent data. We compared the first 10 countries to report 10 SARS-COV-2 cases according to the World Health Organisation (3) to the top 10 invasion rankings predicted by our model. In this validation step we used model simulations that were seeded in China in January 2020 and used flight data for the period January-December 2020.

### 2. Code and data availability

All code used in this analysis is available at https://github.com/j-wardle/flight_passenger_paper. Flight passenger data used in the analysis can be purchased from https://www.iata.org.

### Supplementary Tables and Figures

| Flight scenario | Epidemic centre | Flight data used in "historical" simulations | Flight data used in "contemporary" simulations |
|---|---|---|---|
| MERS | South Korea | June 2014 – May 2015 | June 2015 – May 2016 |
| Zika | Brazil | Mar 2015 – Feb 2016 | Mar 2016 – Feb 2017 |
| SARS-CoV-2 | China | Feb 2019 – Jan 2020 | Feb 2020 – Jan 2021 |

**Table S1. Summary of the time periods that were defined as "*contemporary*" and "*historical*" for the epidemic simulations.** "*Contemporary*" describes flight data that included the period where there was disruption due to an epidemic. "*Historical*" describes flight data from the 12-month period prior to the start of the contemporary period. The epidemic centre refers to the country where the epidemic simulations began.

Please note the deliberate choice for the contemporary periods to not directly align with the true dates of the outbreak. For example, the MERS outbreak in South Korea took place from May to July 2015, the Zika epidemic in Brazil was declared a Public Health Emergency of International Concern in February 2016, and the first cases of SARS-CoV-2 were reported at the very end of December 2019. We chose for the contemporary simulations to begin one month into each of these periods, as this resulted in the contemporary periods containing increased levels of passenger disruption. Consequently, this improved our ability to detect any impacts of using historical data in the models.

| Natural history scenario | R0 | Mean pre-infectious period (days) | Mean infectious period (days) | Mean generation time (days) |
|---|---|---|---|---|
| ‘MERS’-like | 1.2 | 6 | 3 | 9 |
| ‘Zika’-like | 2.0 | 14 | 7 | 21 |
| ‘SARS-CoV-2’-like | 3.0 | 4 | 4 | 8 |

**Table S2. Natural history parameters used in simulation scenarios.** These scenarios were chosen to explore epidemic simulations for pathogens with differing characteristics. Although they were initially informed by the natural history parameter values of MERS, Zika and SARS-CoV-2 from the literature, some adjustments have been made to the parameters we used so that we could explore a range of transmissibility scenarios.

| Country name | Order in which country reported 10 cumulative cases (WHO case database) | Order in which country experienced 10 cumulative infections in simulated epidemics using… | |
|---|---|---|---|
| | | …contemporary movement data | …historical movement data |
| Thailand | 1st | 1st | 1st |
| Singapore | 2nd | 7th | 6th |
| Japan | 3rd | 2nd | 2nd |
| South Korea | 4th | 3rd | 3rd |
| Germany | 5th | 15th | 16th |
| Australia | 6th | 6th | 7th |
| USA | 7th | 4th | 4th |
| Malaysia | 8th | 5th | 5th |
| United Kingdom | 9th | 14th | 12th |
| Vietnam | 10th | 12th | 9th |

**Table S3. Validation of global epidemic model using SARS-CoV-2 data.** The table presents a comparison of the first 10 countries to report 10 cumulative cases of SARS-CoV-2 (3) and the order in which those countries experienced 10 cumulative infections in SARS-CoV-2 epidemic simulations using contemporary or historical movement data. The simulation period for model validation ran from January-December to provide a more suitable comparison to the WHO data.

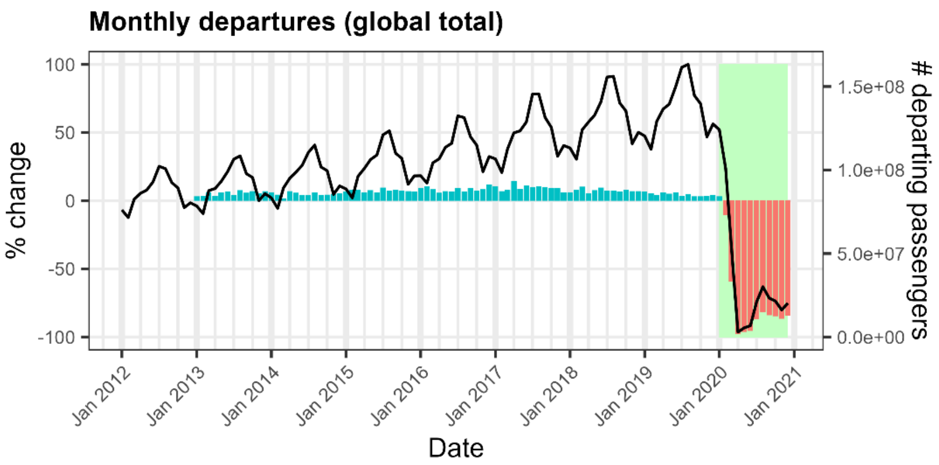


**Figure S1. Changes in departing flight passenger volumes over time (global totals).** The lines show the monthly global numbers of flight passengers (right-hand axis) from January 2012 to December 2020. These numbers exclude passengers taking internal flights (i.e. those that depart and land in the same country). The bars denote the percentage change in flight passenger numbers (left-hand axis) relative to the same calendar month in the previous year. Blue bars represent an increase in passenger numbers, red bars represent a decrease. Green rectangle represents the SARS-CoV-2 pandemic.

A)

B)

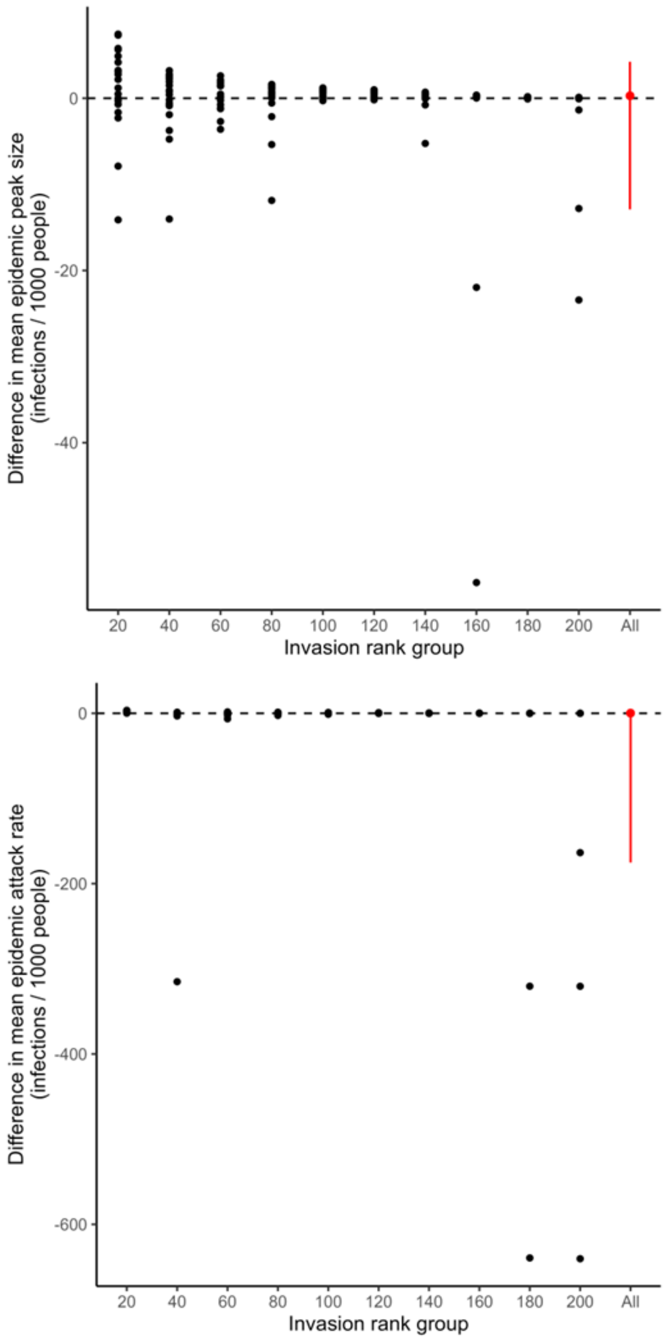


**Figure S2. Difference in A) peak sizes and B) attack rates for SARS-COV-2 epidemics simulated with contemporary vs historical flight data.** Each black dot represents an individual country. Countries are grouped by the average order in which they are invaded (invasion rank) in simulations using contemporary flight data. The red dot summarises across all countries, with the error bar showing the 2.5% and 97.5% quantiles. The difference on the y-axis is obtained by subtracting the mean peak size or attack rate across simulations using historical flight data from the mean values in simulations using contemporary data.

### References


1. International Air Transport Association (IATA). Flight passenger data available from: https://iata.org.
2. World Bank Group. Population Estimates and Projections. Available at: https://datacatalogworldbankorg/dataset/population-estimates-and-projections. 2022.
3. World Health Organisation. WHO Coronavirus (COVID-19) dashboard. Available at: https://data.who.int/dashboards/covid19/about. 2023.
